# High risk landscapes of Japanese encephalitis virus outbreaks in India converge on wetlands, rainfed agriculture, wild Ardeidae, and domestic pigs

**DOI:** 10.1101/2021.09.07.21263238

**Authors:** Michael G. Walsh, Amrita Pattanaik, Navya Vyas, Deepak Saxena, Cameron Webb, Shailendra Sawleshwarkar, Chiranjay Mukhopadhyay

**Author notes:** Address correspondence to: Michael Walsh, PhD, MPH, Senior Lecturer, Infectious Diseases Epidemiology, Marie Bashir Institute for Infectious Diseases and Biosecurity, Sydney School of Public Health, The University of Sydney, 176 Hawksbury Road, Westmead NSW 2145 Australia.

## Abstract

Japanese encephalitis constitutes a significant burden of disease across Asia, particularly in India, with high mortality in children. This zoonotic mosquito-borne disease is caused by the *Flavivirus*, Japanese encephalitis virus (JEV), and circulates in wild ardeid bird and domestic pig reservoirs both of which generate sufficiently high viremias to infect vector mosquitoes, which can then subsequently infect humans. The landscapes of these hosts, particularly in the context of anthropogenic ecotones and resulting wildlife-livestock interfaces, are poorly understood and thus significant knowledge gaps in the epidemiology and infection ecology of JEV persist, which impede optimal control and prevention of outbreaks. The current study investigated the landscape epidemiology of JEV outbreaks in India over the period 2010 to 2020 based on national human disease surveillance data. Outbreaks were modelled as an inhomogeneous Poisson point process. Outbreak risk was strongly associated with the habitat suitability of ardeid birds and pig density, and shared landscapes between fragmented rainfed agriculture and both river and freshwater marsh wetlands. Moreover, risk scaled with Ardeidae habitat suitability, but was consistent across scale with respect to pig density and rainfed agriculture-wetland mosaics. The results from this work provide a more complete understanding of the landscape epidemiology and infection ecology of JEV in India and suggest important priorities for control and prevention across fragmented terrain comprised of wildlife-livestock interface that favours spillover to humans.

## Introduction

Japanese encephalitis virus (JEV) is one of the most substantial causes of childhood encephalitis in Asia(1). While most infections are asymptomatic or mild (approximately 1 in 250 infections present with severe clinical disease), mortality is high among those presenting with encephalitis(1). In India, a country with a high burden of disease caused by JEV, 13.7% of 63,854 acute encephalitis cases from 2010 to 2017 were due to JEV and over 17% of these cases died(2). Although the annual occurrence of Japanese encephalitis (JE) is high, there is considerable heterogeneity in its occurrence across the country with the northeast being a perennial hotspot for outbreaks, although additional far-removed areas of intractable endemicity also persist(2). Japanese encephalitis virus is a mosquito-borne zoonotic Flavivirus with enzootic and endemic transmission in animal and human hosts, respectively, although such baseline transmission is regularly punctuated with more substantial outbreaks(3,4). Outbreaks in India are generally seasonal following monsoon flooding, but transmission can and does happen at any time of the year with rural populations typically at highest risk although some urban locations also experience outbreaks(2).

The infection ecology of JEV is complex and incompletely understood in many landscapes. As a result, viral transmission is often poorly controlled. Culex tritaeniorhynchus is the most important vector for JEV across Asia(4,5) and has a wide distribution in India(5,6). In addition to this highly efficient vector, there are at least four other important vectors (*Cx. vishnui, Cx. gelidus, Cx. fuscocephala*, and *Cx. pseudovishnui*) that also exhibit wide distribution across South and Northeast India(6,7). Given the wide range of suitable habitats for these mosquitoes, exposure to JEV vectors is extensive throughout the country. Wading bird species in the Ardeidae family are the primary reservoirs and maintenance hosts for JEV(8–11), while domestic pigs are key amplifying hosts that frequently accelerate spillover to humans(12–17). This general distinction between host groups notwithstanding, high viremias have been shown in several Ardeidae species, so these maintenance hosts may also simultaneously act as amplifying hosts depending on the nature of their interface with humans or pigs(4). Moreover, some heron species can readily adapt to some agricultural practices (e.g. rice paddies), increasing contact between people and domestic animals in these settings(18). Interestingly, specific maintenance host-mosquito vector-amplifying host interactions have been identified showing *Cx. tritaeniorhynchus* zoophilic feeding preferences for herons and domestic pigs, which may further highlight the importance of interface and the potential for an efficient bridge to human spillover in landscapes that favour these interactions(4).

The biotic factors described above define the vectors and hosts in which JEV circulates and the nature of interspecies interaction that may drive viral transmission dynamics in hosts, but there are equally important abiotic factors that can influence JEV transmission such as wetlands and rainfed agriculture. Heterogenous wetlands not only provide a spectrum of favourable habitat for vectors, they also demarcate critical habitat for key ardeid reservoirs(19). Rainfed agricultural mosaics tend to comprise agricultural systems that 1) are engaged by poorer, subsistence communities, and 2) exhibit far less control of water distribution in the landscape(20). Both wetland habitat and rainfed mosaics can influence mosquito habitats by way of their distribution of water in the landscape, and, since both landscapes can be sensitive to the modulating effects of climate, these could represent important vector foci(21). Moreover, rainfed crop mosaics that lie within or adjacent to wetland habitat may present ecotones of particular risk since these often also present landscapes occupied by key animal hosts and may therefore exhibit multiplicities of JEV transmission (Figure 1). In India, while some states have been recognised as hotspots of annual JEV outbreaks, the landscape epidemiology of JEV has not been thoroughly described in these and other areas of occurrence. The heterogeneity of risk is particularly noteworthy since viable mosquito vectors can be found in most parts of the country. As such, the delineation of JEV outbreak risk across India requires a broader consideration of diverse landscapes, which represent mosaics of wetland habitat, rainfed agriculture, and animal hosts (Figure 1).

**Figure 1.**
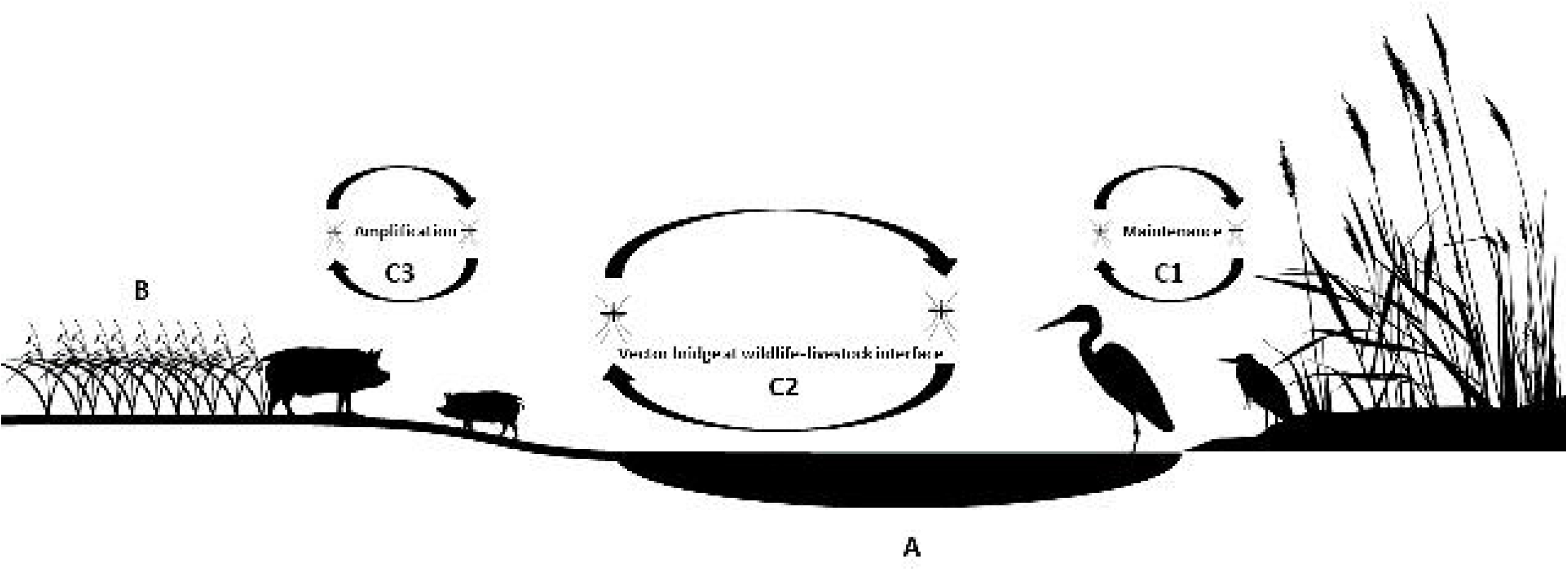
Theoretical representation of landscapes with wetland (A) and rainfed crop (B) mosaics and their potential animal host occupants. Multiple transmission cycles of Japanese encephalitis virus (JEV) may be realised in such landscapes such as transmission among Ardeidae maintenance hosts (C1), shared transmission between ardeid birds and domestic pigs at the wildlife-livestock interface (C2), and concentrated transmission among porcine amplification hosts (C3).

Finally, a complete understanding of the epidemiology and infection ecology of JEV requires an understanding of the differential scaling of biotic and abiotic drivers of JEV activity across landscapes. There is some evidence to suggest that biotic interactions between organisms, and abiotic interactions between organisms and their environment (or environmental filtering), may scale differently with respect to some ecological relationships, particularly with respect to pathogen transmission (22). Under this framework, biotic interactions may dominate at more local scale, whereas abiotic interactions can exhibit greater influence at broader scale. However, there is considerable variation with respect to such phenomena, wherein some relationships may show consistency across scale or may show the reverse between biotic and abiotic feature dominance at scale(23). The scaling of risk is an added dimension of JEV epidemiology that has gone unexplored and represents a further critical knowledge gap. Because the scaling of associations may have consequences for epidemiological and ecological inference, or the scale of interventions that may be deployed as a result, it is necessary to examine how associations between biotic and abiotic features may scale differently.

The current study sought to identify the key landscape features of JEV outbreaks in India. In particular, this investigation examined the associations between JEV occurrence in humans and the distribution of maintenance and amplifying animal hosts, wetland hydrogeography and flow dynamics, rainfed agriculture, and climate. These associations were further interrogated at both local and broad scale to determine whether JEV risk scales differently for biotic and abiotic landscape features. While considerable heterogeneity in risk was anticipated, it was hypothesised that river wetlands and rainfed agriculture with high pig density and high Ardeidae suitability would drive the landscape epidemiology of JEV.

## Methods

### Data sources

The National Centre for Disease Control’s Integrated Disease Surveillance Programme (IDSP) maintains ongoing surveillance of JEV infections under the administration of India’s Ministry of Health and Family Welfare(24). There were 294 laboratory-confirmed and location-unique outbreaks of JEV reported at village level (spatial resolution of 1 arc minute, or approximately 2 km) to the IDSP between 1 January, 2010 and 31 December, 2020. These were included as the primary training data in the current study. As a test of the external validity of these surveillance data, a secondary dataset (n = 27) comprising all available independent, laboratory-confirmed community surveys of human and mosquito infection conducted within the same time period as the IDSP surveillance and with published location data were used to test the performance of models trained with the IDSP surveillance data(25–28).

JE infections often disproportionally affect communities of lower socioeconomic status with limited access to health care, so this study adjusted for potential reporting bias of JEV infections using the distribution of health system performance as a representation of the local capacity to detect cases (see modelling description below). The infant mortality ratio (IMR) was chosen as a proxy for health system performance since it has been validated as representative of health infrastructure and health system performance and used to assess health service delivery and performance in diverse settings(29,30). Moreover, the IMR is correlated with the Inequality-Adjusted Human Development Index (IHDI) and the Human Development Index (HDI) and is therefore an important representation of the economic, social, and environmental structural determinants of population health(29,31). The raster of the IMR was obtained from the Socioeconomic Data and Applications Center (SEDAC) repository(32). Human population density was derived from the Global Rural-Urban Mapping Project estimates for the 2010 population(33) to represent the baseline population at the beginning of the period under study. The raster data product was obtained from the SEDAC repository.

The Global Biodiversity Information Facility (GBIF) was used to acquire all observations of Ardeidae species (241,784 individual observations of 15 species) between 1 January 2010 and 31 December 2020 across India so each species’ distribution could be modelled(34). Pig density data were obtained from the Gridded Livestock of the World(35) (GLW). While pigs been have been demonstrated as key amplifying hosts for JEV, some evidence suggests that poultry may also act as bridging hosts to human spillover in some settings(11,36,37), so we additionally included poultry density in these analyses obtained from the same GLW source. For some regions of the world these data demonstrate non-negligible spatial heterogeneity in error. However in India, the estimates were adjusted by animal censuses at the 2nd and 3rd stage administration levels, corresponding to the district and taluk, respectively, which represented a high level of data verification at a sub-state scale(35).

Due to potential differential accessibility, the background points used to model Ardeidae species distributions were weighted by the human footprint (HFP) (see modelling description below) to correct for potential spatial reporting bias in the observations of these birds. Human footprint raster data were acquired from the SEDAC registry(38) and quantified according to a 2-stage classification system(39). First, a metric for human influence was constructed based on the following eight categories: (1) population density, (2) road proximity, (3) rail line proximity, (4) navigable river poximity, (5) coastline proximity, (6) artificial light at night, (7) rural versus urban location, and (8) land cover. These categories were scored and summed to generate the human influence index (HII), which ranges from 0 (absence of human impact) to 64 (maximum human impact). The ratio of the range of minimum and maximum HII in the local terrestrial biome to the range of minimum and maximum HII across all biomes, expressed as a percentage, is then calculated to produce the HFP metric(39).

The structure of water movement through the landscape was quantified using hydrological flow accumulation obtained from the Hydrological Data and Maps based on SHuttle Elevation Derivatives at multiple Scales (HydroSHEDS) information system (https://hydrosheds.cr.usgs.gov/), which is derived from elevation data of the Shuttle Radar Topography Mission(40). Hydrological flow accumulation measures the quantity of upland area draining into each 500 × 500 m area.

Wetlands were classified using the surface water data from the Global Lakes and Wetlands Database(41). Wetland types represented in the current study comprised: coastal wetland, river, controlled water reservoir, lake, freshwater marsh, swamp, or intermittent wetland(42). To quantify proximity to each wetland type, the proximity function in the QGIS geographic information system was used to create distance rasters for each wetland class(43). The pixel values of these rasters represent the distance in kilometres between each wetland type and all other pixels within the geographic extent under study. The distance rasters were then used to investigate the relationships between distinct wetland environments and JEV outbreaks.

Agriculture data were obtained from the Global Food Security Support Analysis Data (GFSAD) project to describe the geographic extent of crops that employ rainfed water distribution systems at a resolution of 30 arc seconds(44). Two primary classes of rainfed agricultural systems were represented: dominant rainfed crops and fragmented rainfed crop mosaics. A third class, highly fragmented rainfed crop mosaics, was also available but was highly correlated and exhibited considerable overlap with fragmented mosaics and yielded very similar relationships, so this class was considered redundant and not included in this investigation. As with the wetland classes described above, distance rasters were created for both the dominant rainfed crop and fragmented rainfed crop classes using the proximity function in QGIS.

Climate data were obtained from the WorldClim Global Climate database(45). This investigation examined seasonal measures of precipitation due to the distinct seasonal pattern in JEV outbreaks, as particularly marked by monsoon-associated precipitation. Accordingly, rasters for the mean driest quarter precipitation and wettest quarter precipitation, as well as the mean annual temperature, were used in this analysis.

### Statistical Analyses

#### Ardeidae species distribution modelling

An ensemble approach comprising both boosted regression trees (BRT) and random forests (RF) models was used to estimate the landscape suitability of each of the 15 Ardeidae species. Species distribution models (SDMs) based on these machine learning frameworks partition the data space according to algorithms that optimise homogeneity among predictors and a response (e.g. species presence), whereby optimised decision trees are iteratively determined and can capture complex interactions between predictors(46–49). Each SDM under the two distinct modelling frameworks (BRT and RF) was fit using five-fold cross-validation. To prevent artificial spatial clustering of observation data, the data were thinned to include only one observation per pixel in the analysis (S1 Table 1). Mean annual precipitation, mean annual temperature, isothermality, and proximity to surface water comprised the environmental features included in the SDMs. These variables exhibited low correlation with each other (all Pearson’s r < 0.5) and therefore their inclusion together in the models was justified. Model performance, based on the area under the receiver operating characteristic curve (AUC), and model fit, based on the deviance, were used to evaluate each of the two SDM frameworks (BRT and RF) for each ardeid species. Subsequently, an ensemble landscape suitability was estimated for each species from the two SDM frameworks using their weighted mean, with weights based on AUC(50). Potential spatial sampling bias in the GBIF database was adjusted for by sampling background points proportional to the human footprint as a proxy for landscape accessibility. The landscape suitability for each species was modelled at a spatial resolution of 30 arc seconds (∼1 km). Individual species are presented with their number of field observations (and thinned analytical observations) and model metrics in S1 Table 1.

After modelling the distributions of individual Ardeidae species’ landscape suitability, a composite of ardeid suitability was calculated based on the mean of all individual species suitability distributions. The degree of niche overlap(51) between each individual species landscape suitability and the composite species suitability was evaluated to determine the extent of heterogeneity between the species-specific environmental niches. More specifically, niche overlap was assessed to determine (1) if heterogeneity was too extensive to justify a composite representation of Ardeidae species landscape suitability, (2) if a small number of ardeid species demonstrating divergent landscape suitability should be considered individually in concert with a composite representation of landscape suitability for the remaining species, or (3) if the species demonstrated sufficient overlap in their environmental niches to justify a composite representation of Ardeidae species landscape suitability alone. The sdm package(50) in the R platform(52) was used for fitting each model and the derivation of the two-model ensembles to each species and the dismo package was used to compare niche overlap(53).

#### JEV outbreak modelling

The JEV outbreaks were fitted as a point process using homogeneous and inhomogeneous Poisson models(54). This framework allows for the assessment of spatial dependencies among the outbreaks and, where such dependencies are identified, these can be evaluated with respect to environmental features that may account for the observed dependencies.

First, JEV outbreaks were fitted as a homogeneous Poisson process, with conditional intensity,

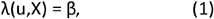

where u designates the geographic locations of outbreaks, X, and β represents the intensity parameter. Intensity is defined as the number of points in a subregion of a defined geographic extent. The homogeneous Poisson model is the null model representing complete spatial randomness (CSR). Under CSR, the expected intensity is proportional to the area of the subregion under consideration(54), i.e., there is no spatial dependency.

Second, the model with the assumption of CSR was compared to an inhomogeneous Poisson process, which incorporates spatial dependency of the outcome (JEV outbreaks) into the model structure and has conditional intensity,

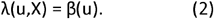

With this model, the intensity is represented as a function of the location, u, of the JEV outbreaks. The inhomogeneous Poisson model supported substantive spatial dependency in JEV outbreak intensity as this was a markedly better fit than the CSR model and also demonstrated significant divergence from CSR in the K-function (see results below). Given the identified spatial dependence in JEV outbreaks, simple and multiple inhomogeneous Poisson models with environmental features were fitted with conditional intensity,

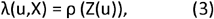

where ρ is the parameter representing the association between the point intensity and the feature Z at location u. The models’ background points were sampled proportional to IMR, as described above, to control for potential reporting bias in the JEV infection surveillance.

As above for the SDMs, the outbreak occurrences were thinned to prevent over-fitting of the models. The data were thinned so that no more than one event was included within each pixel under the two spatial scales investigated at 1.0 and 10.0 arc minutes, respectively (see below). In addition, the environmental covariates were aggregated for these same two spatial scales, 1.0 and 10.0 arc minutes, respectively. Human population density was included in all models as an offset so that the models appropriately represented epidemiological risk. The crude associations between JEV outbreaks and mean dry quarter precipitation, mean wet quarter precipitation, mean annual temperature, hydrological flow accumulation, proximity to each wetland type, proximity to rainfed agricultural systems, the composite landscape suitability of Ardeidae species, pig density, and poultry density were initially assessed individually with a separate simple inhomogeneous Poisson model (S2 Table 2). Features demonstrating bivariate associations with confidence intervals that did not include 0 were included as covariates in the multiple inhomogeneous Poisson models (S3 Figure 1, S4 Figure 2, S5 Figure 3). The features included as covariates in the multiple inhomogeneous Poisson models demonstrated low correlation (all values of the Pearson’s r were <0.5) and so were deemed appropriate to be included together in the models. The associations between JEV outbreaks and landscape features were represented by relative risks, which were computed from the regression coefficients of the inhomogeneous Poisson models. Interaction between fragmented rainfed agriculture and the two dominant wetland types, river and freshwater marsh, were examined separately using a river-rainfed crops model and a freshwater marsh-rainfed crops model with a corresponding interaction term included in each model, respectively. In this way, the interaction between fragmented rainfed agriculture and both river and freshwater marsh wetlands was used to evaluate the impact of their shared landscape mosaics on JEV risk. The Akaike information criterion (AIC) assessed model fit, while the AUC assessed model performance. Importantly, model performance was tested against an independent, laboratory-confirmed dataset derived from the community-based surveys described above. The use of independent data for testing model performance provides a test of the external validity of the results thereby improving model assessment considerably. Model selection was based on a comparison the fit (based on AIC) of the full model to reduced model groups nested on three broad environmental domains (hydrogeography, animal hosts, and climate). These were also compared against a stepwise selection procedure with the full point process model to see if there was any divergence in model selection(55,56). Assessment of K-functions fitted to the JEV outbreaks before and after point process modelling with the specified environmental features was used to determine if these features adequately accounted for the observed spatial dependencies.

As described above, the influence of biotic and abiotic features have been shown to scale differently for some pathogen systems(22). To account for such scaling in the current investigation, models were fitted and assessed at local (1.0 arc minutes) and broad (10.0 arc minutes) scales. Likewise, model fit (AIC) and performance (AUC) were also compared across scales. The R statistical software version 3.6.1 was used to perform the analyses(52). Point process models were fitted and K-functions estimated using the spatstat package(55,56). The silhouette images of Ardeidae, pigs, mosquitoes, and rice in Figure 1 were acquired from phylopic.org and used under Creative Commons license.

## Results

The landscape suitability of individual Ardeidae species demonstrated a high degree of overlap with the composite landscape suitability (niche overlap > 88% for all species, and > 96% for all but one species; S1 Table 1), so the composite measure of Ardeidae suitability was used in the modelling of JEV outbreaks.

At 1.0 arc minute (∼ 2 km), the best fitting and performing models of JEV outbreak risk under fragmented rainfed mosaics with freshwater marsh and fragmented rainfed mosaics with river were the reduced models 7 and 8, respectively, which excluded poultry (S6 Table S3; Table 1). These final models were further supported by the stepwise selection procedure implemented with the point process models. Japanese encephalitis virus outbreaks were strongly associated with both Ardeidae suitability (Table 1 Model 1 - RR = 1.14, 95%C.I. 1.07 – 1.21; Model 2 - RR = 1.13, 95%C.I. 1.06 – 1.21) and pig density (Model 1 - RR = 1.41, 95%C.I. 1.33 – 1.51; Model 2 - RR = 1.40, 95%C.I. 1.31 – 1.49), whereby an increasing presence of both in the landscape was associated with increased risk. Proximity to fragmented rainfed agricultural mosaics (Table 1, Model 1 - RR = 0.978 and Model 2 - RR = 0.979) was associated with increased risk of JEV outbreaks (inverse associations indicate increasing distance from this feature was associated with decreasing risk and vice versa), but not proximity to major non-fragmented rainfed agricultural systems (S2 Table 2). Importantly, proximity to both river and freshwater marsh wetlands were also strongly associated with increased risk, and each modified the association between JEV outbreaks and fragmented rainfed crop mosaics, such that proximity to the rainfed mosaics was associated with risk only in locations where the mosaics were shared with, or adjacent to, the two wetland habitats (Table 1). As expected, climate, especially precipitation, was also strongly associated with JEV outbreaks. Estimates of the distribution of JEV outbreak risk with 95% confidence limits are presented in Figure 3. The spatial dependency apparent in JEV outbreaks as estimated by the homogenous K-function (left panels, Figure 4) was largely accounted for by the final inhomogeneous Poisson models (right panels, Figure 4).

**Table 1.** Adjusted relative risks and 95% confidence intervals for the associations between Japanese encephalitis virus (JEV) outbreaks and each landscape feature as derived from the best fitting inhomogeneous Poisson models. Each landscape feature is adjusted for all others in each of the two models. Models are at a scale of 1.0 arc minutes (∼2 km).

| Landscape feature | Relative risk | 95% confidence interval | p-value |
| --- | --- | --- | --- |
| *Model 1 – Freshwater marsh-rainfed mosaics interaction* |  |  |  |
| Ardeidae landscape suitability (deciles) | 1.14 | 1.07 – 1.21 | 0.00005 |
| Pig density (deciles) | 1.41 | 1.33 – 1.51 | <0.00001 |
| Distance to river (2 km) | 0.74 | 0.56 – 0.99 | 0.02 |
| Distance to freshwater marsh (2 km) | 0.71 | 0.63 – 0.82 | <0.00001 |
| Distance to fragmented rainfed agriculture | 0.978 | 0.969 – 0.987 | <0.00001 |
| Freshwater marsh:fragmented rainfed agriculture | 1.008 | 1.003 – 1.014 | 0.0007 |
| Mean precipitation during the wettest quarter (10 cm) | 1.008 | 1.006 – 1.009 | <0.00001 |
| Mean precipitation during the driest quarter (10 cm) | 1.14 | 1.08 – 1.19 | <0.00001 |
| Mean annual temperature (Celsius) | 0.90 | 0.85 – 0.96 | 0.002 |
| *Model 2 – River-rainfed mosaics interaction* |  |  |  |
| Ardeidae landscape suitability (deciles) | 1.13 | 1.06 – 1.21 | 0.00005 |
| Pig density (deciles) | 1.40 | 1.31 – 1.49 | <0.00001 |
| Distance to river (2 km) | 0.63 | 0.46 – 0.85 | 0.001 |
| Distance to freshwater marsh (2 km) | 0.78 | 0.70 – 0.88 | <0.00001 |
| Distance to fragmented rainfed agriculture | 0.979 | 0.971 – 0.987 | <0.00001 |
| River:fragmented rainfed agriculture | 1.012 | 1.006 – 1.017 | 0.00003 |
| Mean precipitation during the wettest quarter (10 cm) | 1.007 | 1.006 – 1.009 | <0.00001 |
| Mean precipitation during the driest quarter (10 cm) | 1.14 | 1.09 – 1.19 | <0.00001 |
| Mean annual temperature (Celsius) | 0.91 | 0.86 – 0.97 | 0.0009 |

**Figure 2.**
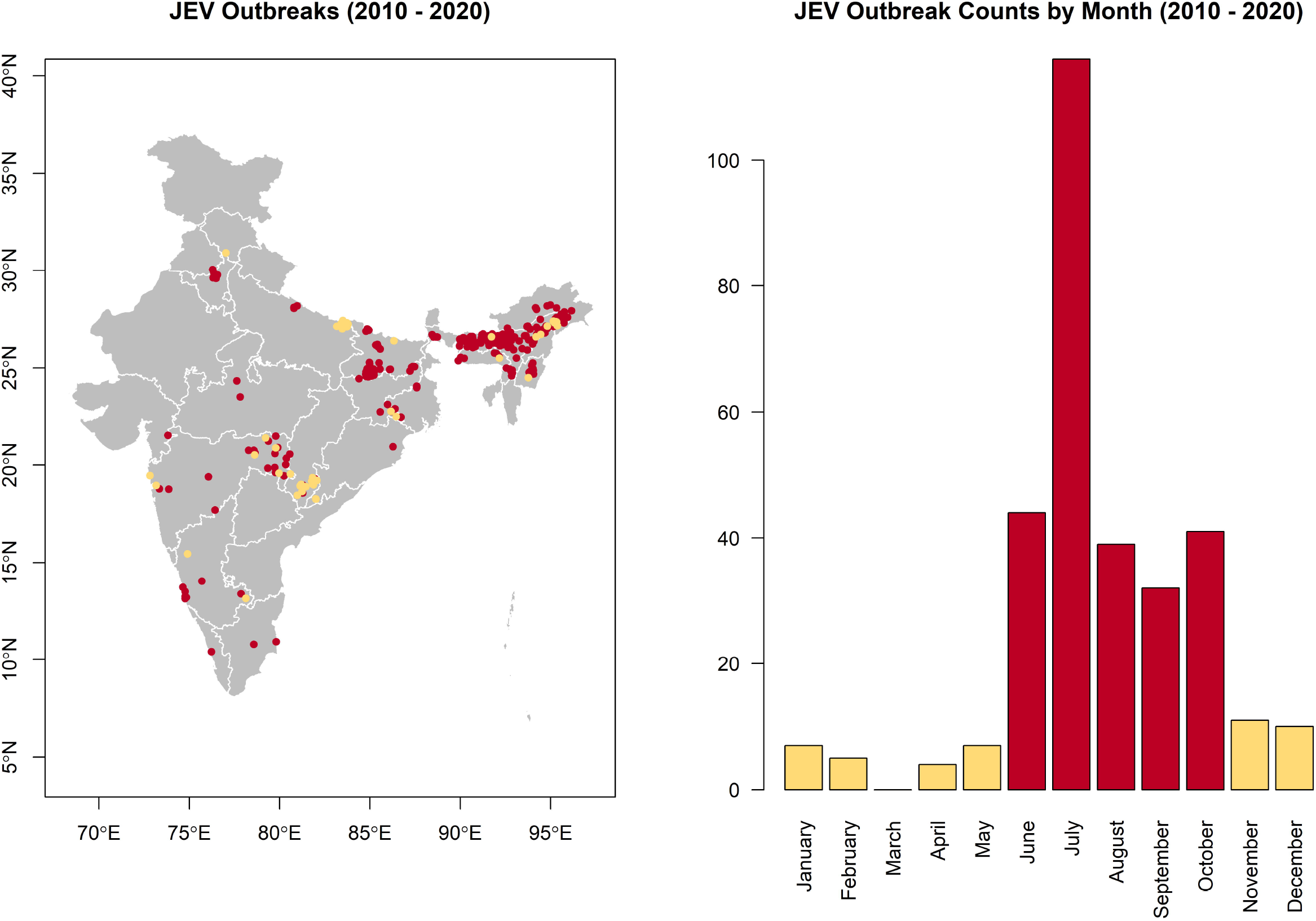
The spatial (left) and temporal (right) distributions of Japanese encephalitis virus (JEV) outbreaks in India. Outbreaks that occurred during the high incidence period are represented in burgundy and those that occurred during the low incidence period in khaki. The map does not reflect the authors’ assertion of territory or borders of any sovereign country including India and is displayed only to present the distribution of JEV occurrence.

**Figure 3.**
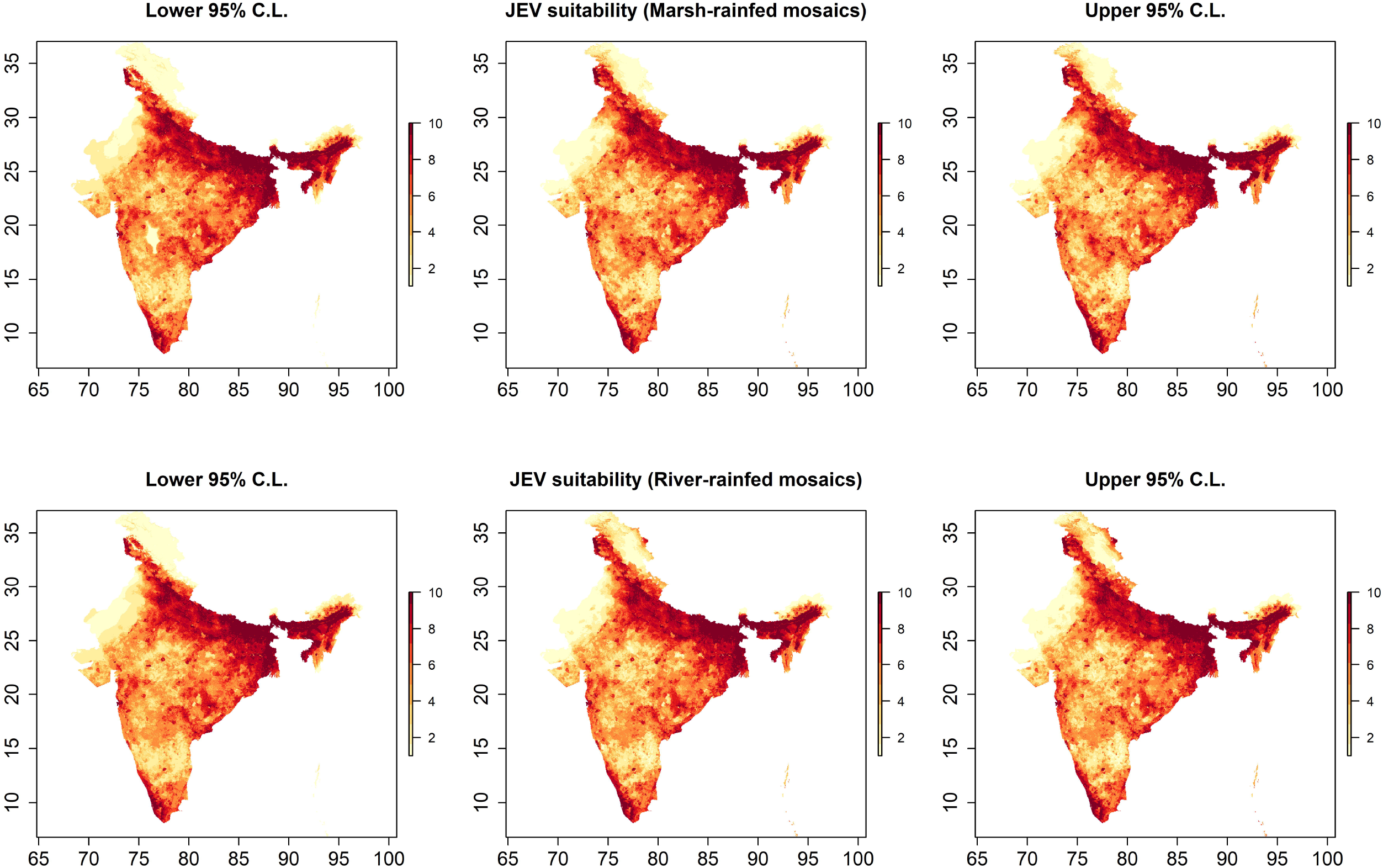
Japanese encephalitis virus (JEV) outbreak risk based on predicted intensity at 1.0 arc minutes (approximately 2 km). The centre panels depict the distribution of JEV risk for freshwater marsh-fragmented rainfed mosaics (top) and for river-fragmented rainfed mosaics (bottom) models as deciles of the predicted intensities from the best fitting and performing inhomogeneous Poisson point process models (Table 1). The left and right panels depict the lower and upper 95% confidence limits, respectively, for the predicted intensities. The map does not reflect the authors’ assertion of territory or borders of any sovereign country including India and is displayed only to present the distribution of JEV occurrence.

**Figure 4.**
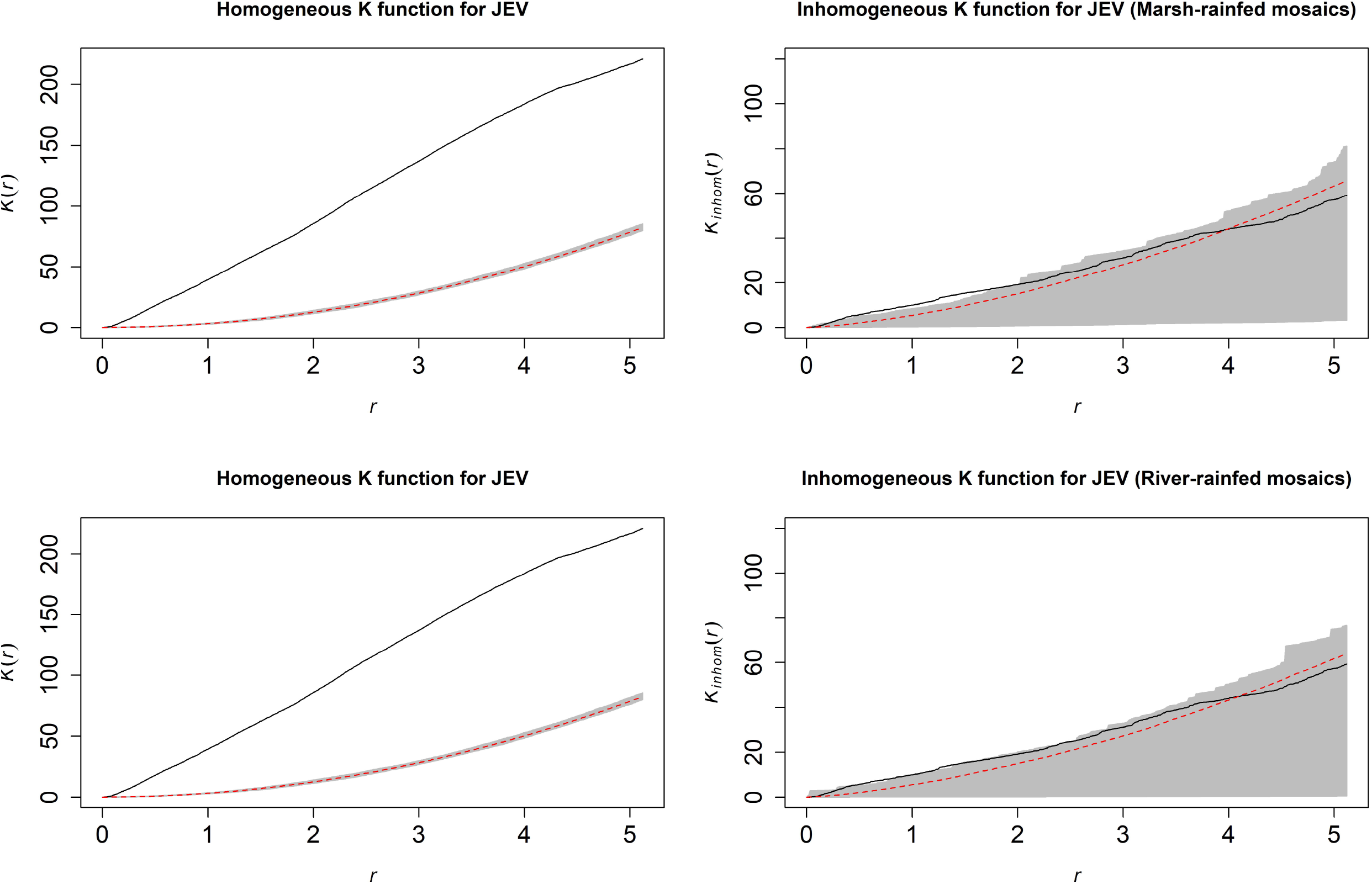
Homogeneous (left panels) and inhomogeneous (right panels) K-functions for the Japanese encephalitis virus (JEV) outbreak point process. The homogeneous K-function is not an appropriate fit due to the spatial dependency in JEV outbreaks as depicted by the divergent empirical (black line) and theoretical functions (the latter is the theoretical function under complete spatial randomness, represented by the dashed red line with confidence bands in grey). In contrast, the freshwater marsh-fragmented rainfed mosaics (top) and river-fragmented rainfed mosaics (bottom) model-based inhomogeneous K-functions show that the spatial dependency was accounted for by the model covariates (overlapping empirical and theoretical functions). The x-axes, r, represent increasing radii of subregions of the window of JEV outbreaks, while the y-axes represent the K-functions.

At 10.0 arc minutes JEV outbreaks continued to demonstrate strong associations with pig density, wetlands and fragmented rainfed mosaics, and precipitation, however Ardeidae suitability and temperature did not continue to manifest influence at this broad scale (S7 Table 4).

## Discussion

This is the first investigation of JEV outbreaks to consider the impact of shared landscapes with key wildlife and domesticated animal reservoirs for JEV, while simultaneously assessing the convergence of natural wetland habitat with rainfed agriculture. Moreover, these landscape features were evaluated at multiple scales to determine whether their influence manifested differently at local and broad scales. Both wild ardeid and domestic pig hosts were strongly associated with JEV outbreak risk at local scale. However, at broad scale only pigs continued to manifest a broad footprint in the landscape. River and freshwater marsh systems and their shared landscapes with fragmented rainfed agriculture were also strongly associated with outbreak risk at both local and broad scale. These differences in risk between animal hosts and the environment demonstrate the importance of considering the ways in which biotic and abiotic features, respectively, comprise complex risk landscapes for JEV that vary with scale. Moreover, these could have potentially important policy implications for the control and prevention of outbreaks. For example, factors that dominate locally such as the sharing of space between ardeid birds and domestic pigs might be ideally targeted for intervention by local municipalities, whereas factors that dominate at broader scale, such as veterinary surveillance of pigs or the development of a novel pig vaccination program, may be more effectively targeted and resourced by state or national authorities.

The family Ardeidae comprises the wading birds, herons (including egrets) and bitterns. Ardeid birds have been recognised as key maintenance hosts for JEV(8–11). Domestic pigs, conversely, are important amplification hosts due to the high viremia associated with porcine infection(12–17). Pigs are also important since these are livestock animals and typically occupy space in close proximity to humans, although several heron species, such as the cattle egret, Bubulcus ibis, are also capable of thriving in anthropogenic landscapes(57). Therefore, as expected, both of these wild and domesticated animal hosts were strongly associated with outbreak risk at local scale in the current study. Interestingly, the influence of Ardeidae suitability appeared to manifest only at local scale, whereas pig density was associated with risk at local and broad scale. The difference in scaling of ardeid and pig hosts may reflect differences in the influence of Ardeidae-pig interfaces at different scales, whereby the influence of domestic pigs on JEV outbreaks may manifest regionally beyond the local impact of their interface with ardeid hosts, whereas the influence of the ardeid hosts may be confined to their local interface with pigs. Importantly, the current study did not observe and assess specific interactions between ardeid birds and pigs across India, which precludes any definitive conclusions about the roles of these hosts in the infection ecology of JEV at different scales of influence. Field investigations of interspecific interactions in local settings will be required to verify the results from the current study and ultimately define how different classes of hosts operate with respect to viral circulation and spillover at scale.

Wetlands can provide important habitat for mosquitoes and therefore increased outbreak risk associated with the provision of a stable source of surface water in these habitats is intuitive. Nevertheless, wetland systems are not homogeneous geomorphologically or ecologically, and neither were they homogeneous with respect to JEV occurrence as clearly demonstrated by the lack of association between outbreak risk and proximity to any surface water type (S2 Table 2). Instead, river and freshwater marsh wetlands dominated JEV outbreak risk, with both also demonstrating interaction with fragmented rainfed agriculture suggesting that shared landscapes of wetland habitat and fragmented rainfed mosaics may be particularly important to the landscape epidemiology of JEV outbreaks. These associations are intuitive because wetland-rainfed agricultural mosaics may represent landscapes of more seasonally stable precipitation compared to rainfed crops that are far removed from wetland habitat. This was further supported by the strong and independent association with precipitation that was observed at both local and broad scale. Furthermore, fragmented rainfed mosaics within or adjacent to wetlands may also demarcate landscapes with limited control of water dispersal following inundation(20), which is particularly relevant to the annual monsoon flooding and which corresponds to the season of highest JEV incidence. It is also important to note that rainfed agriculture is typically a system employed by resource-limited subsistence farmers, with fragmented agricultural landscapes often corresponding to more economically disadvantaged communities(20), and which also tend to represent a preponderance of the annual JEV incident cases(2). Therefore, not only do these findings provide further insight into the epidemiology of JEV outbreaks, they also identify vulnerable communities that are likely to be at greatest risk and which may yield maximum benefit from targeted resource allocation to prevent future outbreaks.

As expected, increasing precipitation was associated with increased JEV outbreak risk at both local and broad scale. Interestingly, temperature was inversely associated JEV outbreak risk at local scale, but demonstrated no association at broad scale. While JEV outbreaks were widely distributed across India, there was a preponderance of occurrence in the wettest areas of the country, which also tend to coincide with areas of slightly lower mean annual temperature and may reflect this inverse association at local scale. Alternatively, areas of more extreme heat in India may exist within a temperature spectrum that is less favourable to the provision of habitat for reservoirs and vectors, to vector development and longevity, to the extrinsic incubation period in vectors, or the areas of highest temperature regimes may simply be coincident with precipitation and humidity regimes that are less favourable(4). Regardless, these associations are based on climate rather than weather. As such, future work will need to explore the effects of specific weather events and patterns with the requisite temporal resolution to link fluctuations in precipitation and temperature with individual JEV outbreaks. For example, one study examined a long-term time series of JEV occurrence and found that increases in both rainfall and temperature were associated with increased risk(21). However, this work was limited to one district in one state, so more work will require examination across many more of India’s heterogenous landscapes to better understand how weather fluctuation may operate in different landscapes. Nevertheless, the association between JEV and precipitation has shown broad geographical consistency as manifested in China, for example, where cases were mostly concentrated in landscapes with annual precipitation greater than 400 mm irrespective of whether these landscapes were characterised by warm-temperate, semitropical or tropical climate regimes(58).

It is important to acknowledge and discuss some additional limitations attending this work. First, although the national IDSP surveillance system was used to capture all reported outbreaks under investigation, we recognise that reporting bias may still be present. To correct for potential reporting bias, rather than randomly selecting background points for the point process models, background sampling was instead weighted by the distribution of IMR as a robust indicator of health system accessibility and infrastructure. Second, the species distribution models used to construct Ardeidae suitability were based on human observations and so are also subject to reporting bias, insofar as bird accessibility is likely to impact reporting effort. Reporting bias in Ardeidae observations was corrected by weighting the sampling of background points by HFP as an indicator of accessibility. In addition, while this study was able to estimate the landscape suitability of several Ardeidae species, there were some species for which there were too few observations to validly model suitability. As such, we concede that this work is not an exhaustive representation of all possible species niches and therefore some aspects may yet remain undescribed by these findings. Third, the climate measures interrogated in the models presented were based on decadal averages over the period from 1950 to 2000, which assumes homogeneity over this time period as well as over the period of JEV outbreak surveillance under investigation. However, the current study sought to model the influence of climate features in the landscape rather than specific weather events, so these assumptions were deemed appropriate.

This study showed that JEV risk in India was strongly associated with the distribution of animal hosts and the shared landscapes between fragmented rainfed agriculture and river and freshwater marsh wetlands. Importantly, animal hosts operated with different degrees of influence at local and broad scale, which may provide unique opportunities to target distinct aspects of JEV landscape epidemiology with differential resource allocation by local and state (or national) municipalities, respectively, to optimise control and prevention of JEV outbreaks. For example, the World Health Organisation has outlined potential forms of landscape manipulation and modification, such as the rotation or synchronisation of crop cycles, alternating crop varieties with variable growing seasons, or mechanical intervention on water movement through the landscape to subvert vector breeding(59). Moreover, the mitigation of vector abundance directly associated with rainfed agriculture may be negated where wetland habitat is present (i.e. in fragmented mosaics) representing a refuge for mosquitoes from control measures and therefore a need for increased attention. Alternatively, there may be opportunities for the repositioning of livestock animal pens at sites more distal to human residences, or locations of agricultural activity, to limit the vector-animal-human interface(59). These kinds of hyperlocal interventions could be ideally suited to administration by local municipalities such as the sub-district taluks (tehsils), particularly since such interventions often require working closely with affected communities. In contrast, broader interventions such as those involving resource-intensive vaccination campaigns of humans (or livestock) may be more effectively orchestrated at the state or national levels. The findings highlight the importance of developing collaborative surveillance infrastructure for vectors, animals, and humans across scale such that an effective surveillance system will require operation and communication within and between taluks, districts, and states. These surveillance systems should also remain adaptive to the influence of land use change, climate change, and urbanisation that may also influence future risks of JEV in India.

## Data Availability

Data will be made publicly available.

