## Supplementary material for "High risk landscapes of Japanese encephalitis virus outbreaks in India converge on wetlands, rainfed agriculture, wild Ardeidae, and domestic pigs"

S1 Table 1. Ardeidae species niche comparisons based on ensemble species distribution models. Each species listed represents that species' modelled suitability with the associated number of observations of the species in the field (and the number of species used for analysis after thinning in parentheses), model fit (deviance), model performance (area under the receiver operating characteristic curve (AUC)), and individual niche overlap with the composite landscape suitability.

| **Ardeidae species** | **Number of field observations** | **Deviance** | **AUC (%)** | **Niche overlap (%)** |
|---|---|---|---|---|
| *Ardea alba* | 18173 (11794) | 0.83 | 89 | 99.7 |
| *Ardea cinerea* | 17774 (11006) | 0.85 | 89 | 99.4 |
| *Ardea purpurea* | 16406 (9784) | 0.78 | 91 | 99.6 |
| *Ardeola grayii* | 56018 (30755) | 0.87 | 88 | 99.7 |
| *Bubulcus coromandus* | 290 (207) | 0.87 | 88 | 98.3 |
| *Bubulcus ibis* | 53548 (32862) | 0.91 | 87 | 99.6 |
| *Butorides striata* | 3398 (2576) | 0.84 | 89 | 99.5 |
| *Dupetor flavicollis* (*Ixobrychus flavicollis*) | 1602 (1278) | 0.77 | 91 | 98.7 |
| *Egretta garzetta* | 36332 (22326) | 0.87 | 88 | 99.8 |
| *Egretta gularis* | 3081 (2050) | 0.41 | 98 | 96.4 |
| *Egretta intermedia* (*Ardea intermedia*) | 22406 (14473) | 0.82 | 89 | 99.8 |
| *Gorsachius melanolophus* | 112 (107) | 0.54 | 94 | 88.6 |
| *Ixobrychus cinnamomeus* | 2151 (1810) | 0.78 | 91 | 99.1 |
| *Ixobrychus sinensis* | 2300 (1739) | 0.71 | 92 | 98.8 |
| *Nycticorax nycticorax* | 8193 (5473) | 0.86 | 89 | 99.7 |

S2 Table 2. Crude, bivariate regression coefficients and 95% confidence intervals for the associations between Japanese encephalitis virus outbreaks and each landscape feature as derived from an inhomogeneous Poisson model with only the one feature included in the model. Note: Proximity to wetland and rainfed agriculture was the specific focus of these particular landscape features, so only those features of these two classes that demonstrated significant inverse associations were included in the multiple point process models.

| **Landscape feature** | **AIC** | **Coefficient** | **95% confidence interval** | **p-value** |
|---|---|---|---|---|
| Null model | 875.58 | | | |
| **Climate** | | | | |
| Mean dry quarter precipitation (10 mm) | 559.99 | 0.14 | 0.11 – 0.17 | <0.00001 |
| Mean wet quarter precipitation (10 mm) | 550.21 | 0.006 | 0.005 – 0.008 | <0.00001 |
| Mean annual temperature (Celsius) | 583.85 | -0.08 | -0.10 – -0.06 | <0.00001 |
| **Hydrogeography and surface hydrology** | | | | |
| Distance to freshwater marsh (2 km) | 415.16 | -0.67 | -0.79 – -0.55 | <0.00001 |
| Distance to lakes (2 km) | 604.09 | 0.79 | 0.14 – 1.42 | 0.01 |
| Distance to rivers (2 km) | 510.64 | -1.16 | -1.43 – -0.89 | <0.00001 |
| Distance to coastal marsh (2 km) | 585.58 | 0.16 | 0.09 – 0.22 | <0.00001 |
| Distance to any surface water (2 km) | 609.76 | -0.03 | -0.06 – 0.01 | 0.07 |
| Hydrological flow accumulation | 610.83 | 0.00 | -0.00005 – 205597 | 0.22 |
| **Rainfed agricultural systems** | | | | |
| Distance to major rainfed agriculture (2km) | 611.30 | -0.005 | -0.02 – 0.006 | 0.19 |
| Distance to fragmented rainfed agriculture (2km) | 575.70 | -0.016 | -0.022 – -0.010 | <0.00001 |
| **Animal hosts** | | | | |
| Ardeidae landscape suitability (deciles) | 582.83 | 0.13 | 0.08 – 0.18 | <0.00001 |
| Pig density (deciles) | 355.16 | 0.43 | 0.36 – 0.49 | <0.00001 |
| Poultry density (deciles) | 536.77 | 0.22 | 0.18 – 0.27 | <0.00001 |

S3 Figure 1. Climate feature distributions.

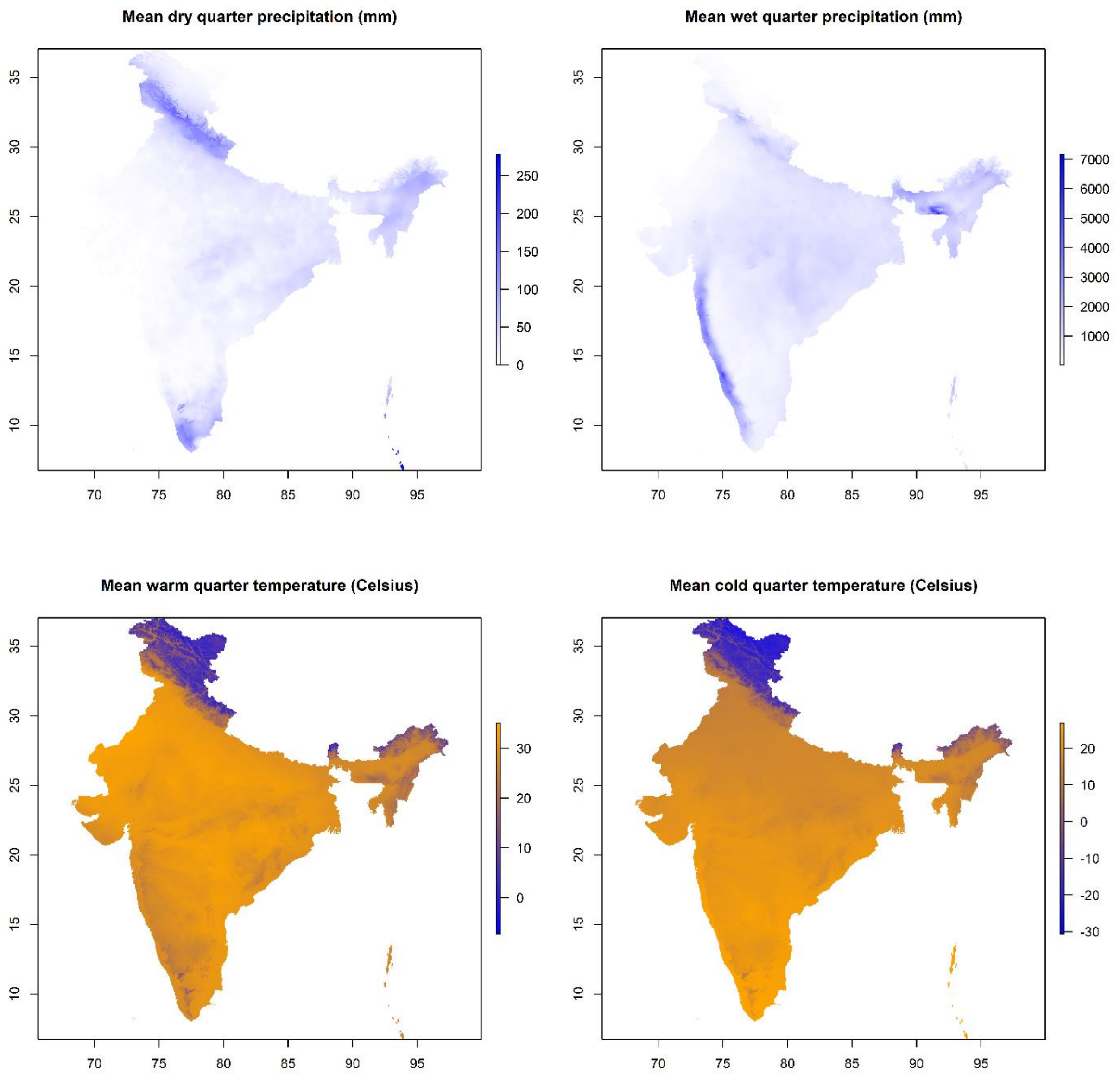

S4 Figure 2. Wetland and rainfed agriculture feature distributions.

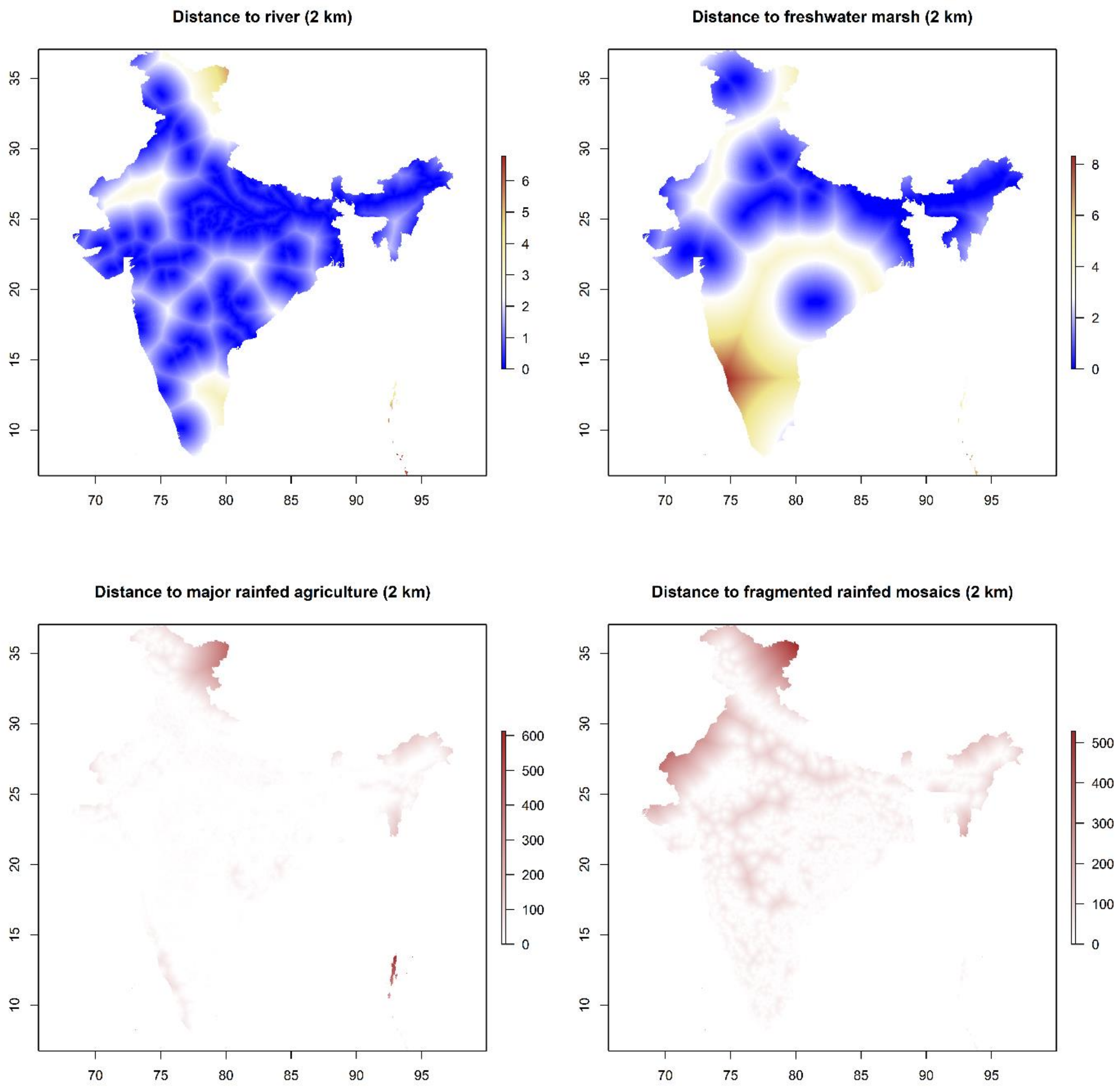

S5 Figure 3. Animal host feature distributions.

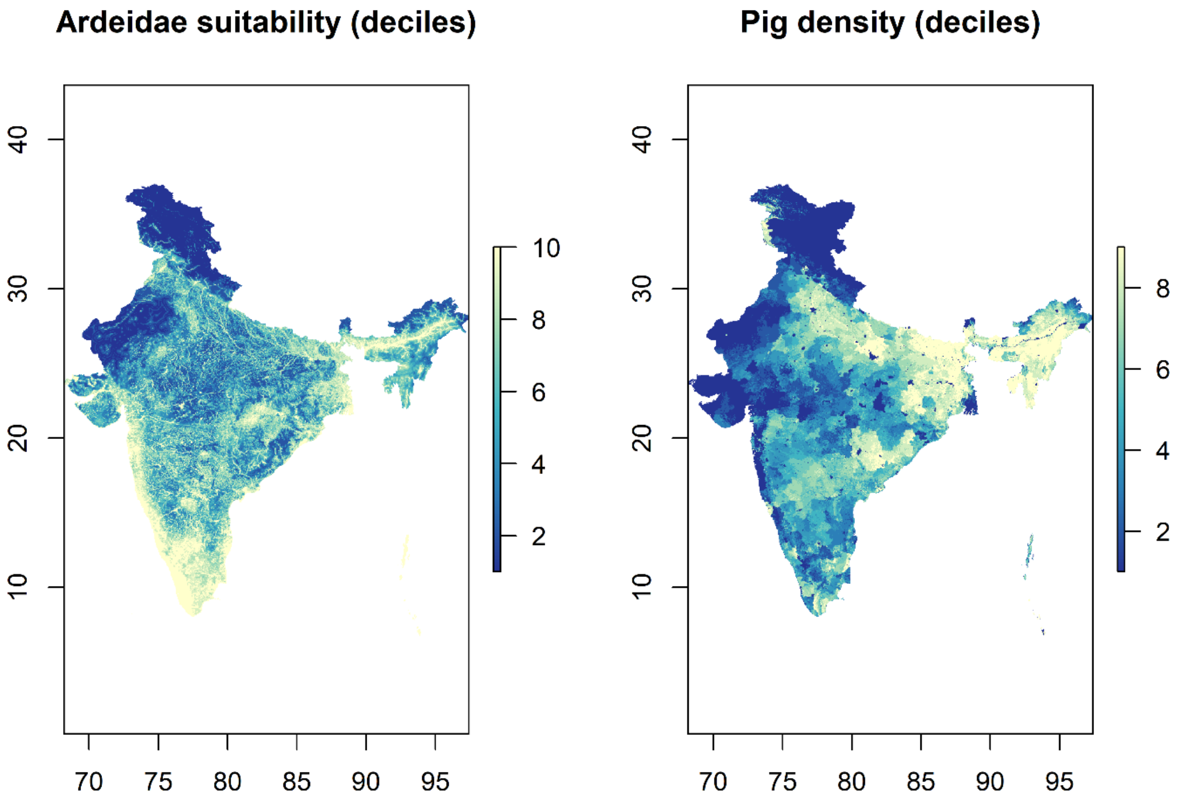

S6 Table 3. Japanese encephalitis virus (JEV) outbreak multiple inhomogeneous Poisson process model comparisons by Akaike information criterion (AIC) and area under the receiver operating characteristic curve (AUC). All models are presented at local (1.0 arc minutes) and broad (10.0 arc minutes) scale. Each nested multiple point process model includes those variables that were bivariately associated with JEV outbreaks (S2 Table 2).

| Pont process models | Scale | AIC | AUC (%) |
|---|---|---|---|
| **Model 1 (Climate only)**: *Mean wet quarter precipitation + mean dry quarter precipitation + mean annual temperature* | | | |
| | 1.0 arc minutes | 503.72 | 89.9 |
| | 10.0 arc minutes | 591.78 | 88.3 |
| **Model 2 (Wetlands only)**: *Freshwater marsh proximity + river proximity* | | | |
| | 1.0 arc minutes | 438.07 | 78.2 |
| | 10.0 arc minutes | 551.18 | 81.5 |
| **Model 3 (Reservoir hosts only)**: *Ardeidae suitability + pig density + poultry density* | | | |
| | 1.0 arc minutes | 295.19 | 91.1 |
| | 10.0 arc minutes | 404.08 | 92.6 |
| **Model 4 (Full, no interaction)**: *Ardeidae suitability + pig density + poultry density + fragmented rainfed ag proximity + freshwater marsh proximity + river proximity + mean wet quarter precipitation + mean dry quarter precipitation + mean annual temperature* | | | |
| | 1.0 arc minutes | 98.49 | 94.4 |
| | 10.0 arc minutes | 292.02 | 95.7 |
| **Model 5 (Full, #1)**: *Ardeidae suitability + pig density + poultry density + fragmented rainfed ag proximity + freshwater marsh proximity + rainfed ag:freshwater marsh interaction + river proximity + mean wet quarter precipitation + mean dry quarter precipitation + mean annual temperature* | | | |
| | 1.0 arc minutes | 92.76 | 93.7 |
| | 10.0 arc minutes | 288.37 | 95.5 |
| **Model 6 (Full, #2)**: *Ardeidae suitability + pig density + poultry density + fragmented rainfed ag proximity + freshwater marsh proximity + river proximity + rainfed ag:river interaction + mean wet quarter precipitation + mean dry quarter precipitation + mean annual temperature* | | | |
| | 1.0 arc minutes | 94.46 | 93.8 |
| | 10.0 arc minutes | 289.36 | 95.3 |
| **Model 7 (Final, #1)**: *Ardeidae suitability + pig density + fragmented rainfed ag proximity + freshwater marsh proximity + rainfed ag: freshwater marsh interaction + river proximity + mean wet quarter precipitation + mean dry quarter precipitation + mean annual temperature* | | | |
| | 1.0 arc minutes | 91.88 | 93.7 |
| | 10.0 arc minutes | 287.91 | 96.1 |
| **Model 8 (Final, #2)**: *Ardeidae suitability + pig density + fragmented rainfed ag proximity + freshwater marsh proximity + river proximity + rainfed ag:river interaction + mean wet quarter precipitation + mean dry quarter precipitation + mean annual temperature* | | | |
| | 1.0 arc minutes | 93.75 | 93.9 |
| | 10.0 arc minutes | 288.98 | 95.5 |

S7 Table 4. Adjusted relative risks and 95% confidence intervals for the associations between Japanese encephalitis virus (JEV) outbreaks and each landscape feature as derived from the best fitting inhomogeneous Poisson models. Each landscape feature is adjusted for all others in each of the two models. Models are at a scale of 10.0 arc minutes (~20 km).

| **Landscape feature** | **Relative risk** | **95% confidence interval** | **p-value** |
|---|---|---|---|
| *Model 1 – Freshwater marsh-rainfed mosaics interaction* | | | |
| Pig density (deciles) | 1.50 | 1.38 – 1.62 | <0.00001 |
| Distance to freshwater marsh (2 km) | 0.76 | 0.68 – 0.85 | <0.00001 |
| Distance to fragmented rainfed agriculture | 0.982 | 0.974 – 0.990 | <0.00001 |
| Freshwater marsh:fragmented rainfed agriculture | 1.003 | 1.002 – 1.005 | 0.00003 |
| Mean precipitation during the wettest quarter (10 cm) | 1.010 | 1.008 – 1.012 | <0.00001 |
| Mean precipitation during the driest quarter (10 cm) | 1.14 | 1.08 – 1.19 | <0.00001 |
| *Model 2 – River-rainfed mosaics interaction* | | | |
| Pig density (deciles) | 1.49 | 1.37 – 1.61 | <0.00001 |
| Distance to river (2 km) | 0.74 | 0.56 – 0.98 | 0.017 |
| Distance to freshwater marsh (2 km) | 0.82 | 0.73 – 0.92 | 0.0003 |
| Distance to fragmented rainfed agriculture | 0.981 | 0.973 – 0.99 | <0.00001 |
| River:fragmented rainfed agriculture | 1.008 | 1.005 – 1.011 | <0.00001 |
| Mean precipitation during the wettest quarter (10 cm) | 1.010 | 1.008 – 1.012 | <0.00001 |
| Mean precipitation during the driest quarter (10 cm) | 1.15 | 1.09 – 1.20 | <0.00001 |
